# Bivariate joint modelling for mixed continuous and binary responses in the presence of non-monotone, non-ignorable missingness: The case of prostate cancer

**DOI:** 10.1101/2023.08.22.23294418

**Authors:** Madiha Liaqat, Shahid Kamal, Florian Fischer

**Author notes:** **Corresponding author:** Dr. Florian Fischer, Charité – Universitätsmedizin Berlin, Institute of Public Health, Charitéplatz 1, 10117 Berlin, Germany.

## Abstract

Joint modelling for mixed longitudinal responses has played a prominent part in disease decision-making. It is based on a joint strategy of estimating joint likelihood with shared random effects. Non-ignorable missingness in outcomes increases complexity in joint model; a shared parameter model is proposed to incorporate non-ignorable missing data for joint modelling of longitudinal responses and missing data mechanism. Parameters are estimated under the Bayesian paradigm and implemented via Markov chain Monte Carlo (MCMC) methods with Gibbs sampler. To demonstrate the effectiveness of the proposed method, the joint model is applied to analyze a prostate cancer dataset. The objective is to assess whether there is an association between two mixed longitudinal biomarkers, which could have important implications for understanding disease progression and guiding treatment decisions. The dataset contains non-monotone missingness pattern. To evaluate the performance and robustness of the proposed joint model, simulation studies are conducted.

## 1. Introduction

Longitudinal studies have a wide range of applications in medicine, where measurements are repeatedly observed for individuals over time. These measurements are correlated even for different types of responses and collected for the same individuals. For example, in oncology research, more than one biomarker of mixed types is measured repeatedly over time for the same patient. These biomarkers are inherently associated – and this association is accounted for valid conclusions. Simultaneous modelling of mixed biomarkers is an optimal approach to account for correlation among multiple biomarkers [1].

Missing data is an unavoidable issue in longitudinal studies because patients may not come for a pre-specified follow-up visit at a particular time point for many reasons. In this regard, missingness exists in collected data and creates hindrances to extracting inference without making assumptions about the missingness mechanism. According to Rubin [2], missing completely at random (MCAR) and missing at random (MAR) mechanisms being not dependent upon unobserved measurements are ignorable. Not missing at random (NMAR) mechanism may depend upon both observed and unobserved responses and is considered non-ignorable. Non-ignorable missingness mechanism is considered in data analysis with longitudinal measurements to get unbiased estimates.

In literature, selection models, pattern-mixture models (PMMs), and shared parameter models (SPMs) are mostly applied to tackle non-ignorable missing data. These models use different factorizations for missingness and measured processes to jointly model longitudinal responses incorporating missingness. The SPM approach is widely applied to model MAR data in longitudinal responses. Still, there exists a gap to employ SPM for NMAR. Non-ignorable missing data is addressed by employing logistic or probit models, which are specifically designed to handle situations where the missing data mechanism is influenced by unobserved variables or responses, resulting in NMAR mechanisms. By integrating the logistic or probit model into the analysis, researchers can effectively mitigate potential biases caused by non-ignorable missing data that consequently makes precise inferences based on the available data. Logistic and probit models provide valuable insights into the underlying patterns of missingness, contributing to obtaining reliable results in longitudinal studies and other scenarios where missing data is common.

Joint modelling terms are applied to simultaneously analyzed more than one outcome. Many methods have been proposed to jointly analyze single or multivariate longitudinal measurements alongside event time data in recent years [3,4]. Another joint modelling term is used to simultaneously analyze more than one longitudinal outcome possibly of different types using random effects, marginal or conditional models [5,6]. Catalano and Ryan [7] analyzed toxicity data proposing bivariate latent variable models to account for the relationship between fetal weight and malformation in live fetuses. Leon and Carriere [8] studied one longitudinal and one binary response to assess maternal smoking’s effect on the respiratory illness of children. Liu et al. [9] formulated a joint model to handle longitudinal binary and continuous responses by incorporating an ignorable missing data mechanism. Li et al. [10] applied joint modelling for continuous, binary, and ordinal responses under the Bayesian framework, while Kürüm et al. [11] proposed a joint model to analyze binary and continuous responses under frequentist statistics.

Under the umbrella of non-ignorable missing data (NMAR) fall both non-monotone and monotone missingness patterns, which in turn provide incomplete observations. Intermittent missingness comes under non-monotone missingness pattern, where an individual misses any visit during the follow-up time, and returns to show up for subsequent visits. The monotone missingness pattern usually refers to informative drop-out, where an individual leaves the study before completion and never comes back to complete follow-up time. Stubbendick and Ibrahim [12] used a likelihood-based approach to incorporate non-monotone NMAR by proposing a joint likelihood of outcome and missing data indicators. Hogan et al. [13] worked on monotone missingness in longitudinal data. Gaskins et al. [14] worked on non-ignorable drop-out mixed longitudinal responses using PMM under joint modelling methodology.

In this article, a joint model is proposed for longitudinal continuous-binary biomarkers using a conditional modelling approach, and two shared parameter models are proposed for non-ignorable missingness under the conditional model. For estimating the model’s parameters different approaches are employed: likelihood-based approach, linear programming technique, and Bayesian approach. We work under the Bayesian framework to obtain parameter estimates.

The article presents a robust statistical approach to analyze longitudinal mixed-effects biomarker measurements, accounting for non-monotone, non-ignorable missingness in the context of prostate cancer research. By utilizing joint modelling techniques and considering the missingness mechanisms, the researchers aim to obtain more reliable and meaningful results from their data analysis.

## 2. Motivation: Prostate cancer dataset

This research is motivated by a comprehensive dataset obtained from prostate cancer patients who underwent external beam radiotherapy (EBRT), Androgen deprivation therapy (ADT), and a combination of ADT along with other therapies. This dataset spans from 2012 to 2019 and includes information collected during up to 5 follow-up visits after treatment. The data was sourced from Mayo hospital, a renowned referral hospital in Pakistan. Data was received in 2021 by the authors of this study. The authors had no access to information that could identify individual participants.

The evaluation of patients took place from the pre-treatment phase to the post-treatment follow-up. During the pre-treatment phase, demographic characteristics and medical history of the patients were recorded. We specifically focused on patients’ age, body mass index (BMI), and Gleason score as baseline predictors for our study. Blood tests were also conducted to measure prostate-specific antigen (PSA), alkaline phosphatase (ALT), platelets, and bilirubin. These biomarkers were tracked after treatment at each follow-up visit, with PSA and ALP values considered as indicators of prostate cancer progression after treatment, and endogenous platelets and bilirubin as time-dependent covariates. Our study included a total of 1,504 patients diagnosed with prostate cancer, out of which 1,026 received ADT and combination therapies as part of their treatment.

## 3. Model specification

### 3.1 Model for longitudinal measurements

Data analysis is described by the joint model specification; sub-models illustrate such mixed types of complex data modelling. The first sub-model assumes to follow linear mixed-effects model specified for continuous longitudinal biomarker. Binary longitudinal response is assumed to follow mixed-effects logistic regression model.

Let *Y*_*ij*_ be the continuous longitudinal outcome for *i*^*th*^ individual *i* = 1,2,3,….,*n* at time *t*_*ij*_ *j* = 1,2,3,….,*m*^*y*^, assume linear mixed effects model as

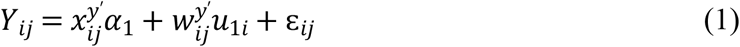

where, *α*_1_ is a *p*^*y*^ dimensional vector of fixed effects regression coefficients, 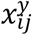 vector of covariates. 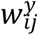 is *q*^*y*^ dimensional vector of random effects, *u*_1*i*_ is a vector of random effects, that is independently and identically distributed as multivariate normal with mean vector 0 and covariance matrix *D*. 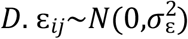 is the random error term.

Let, *Z*_*ij*_ be the binary repeated measurements observe for *i*^*th*^ individual, *i* = 1,2,3,….,*n* at time *s*_*ij*_. Where *s*_*ij*_ = *t*_*ij*_, *j* = 1,2,3,….,*m*^*z*^, binary response *Z*_*ij*_ assumes to follow logistic mixed effects model that is given by

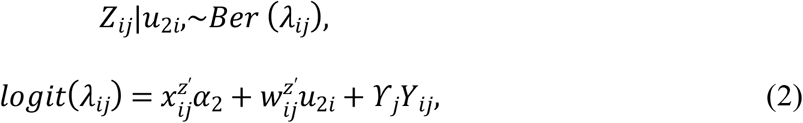

where, *α*_2_ is a *p*^*z*^ dimensional vector of fixed effects regression coefficients, 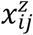 vector of covariates. 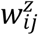is *q*^*y*^ dimensional vector of random effects, *u*_2*i*_∼*MVN*(0, *D*) is a vector of random effects, with the addition of associated parameter *ϒ*_*j*_ to measure the effect of response *Y*_*ij*_ at time *t*_*ij*_ on response *Z*_*ij*_ at the same time *s*_*ij*_.

### 3.2 Missing data mechanism and shared parameter model

Longitudinal data are not fully observed which leads to incomplete measurements. The most important consideration is to specify an appropriate missing data mechanism based on assumptions related to unobserved and observed data. Little and Schluchter [15] as well as Fitzmaurice and Laird [16] applied general location model described by Olkin and Tate [17] assuming the ignorability assumption. This assumption leads to the usage of observed responses only without considering a model for missingness mechanism. Usually, the expectation-maximization (EM) algorithm is employed for parameter estimation.

In this study, we examine two distinct missingness mechanisms to account for the presence of two different missingness patterns in longitudinal outcomes, two SPM are specified for missingness mechanisms.

Let *R*_*i*_ = (*R*_*i*1_, *R*_*i*2_,*R*_*i*3_,….,*R*_*in*_)′ be the vector of response indicators, if *Y*_*ij*_ observed *R*_*ij*_ = 1, and *R*_*ij*_ = 0 for missing *Y*_*ij*_. In this paper, a SPM based on non-ignorable missigness mechanisms for *Y*_*i*_ and *Z*_*i*_ consider, assuming logistic mixed effects regression models for 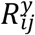 and 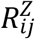 responses.

Let 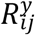 be the missingness indicator for continuous response at time *t*_*ij*_, such that 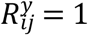 if *Y*_*ij*_ is not fully observed, the model for 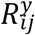 is given by

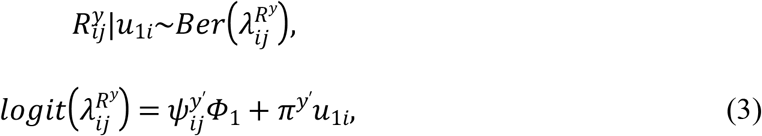

Let 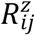 be the missingness indicator for continuous response at time *s*_*ij*_, such that 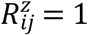 if *Z*_*ij*_ is not fully observed, the model for 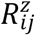 is given by

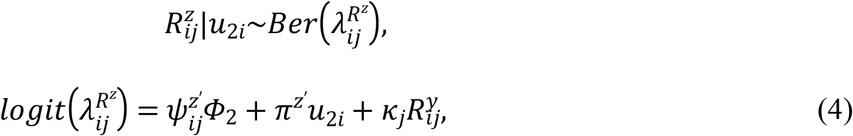

To formulate joint distribution of responses given the random effects, let *Y*_*ij*_ and *Z*_*ij*_ are partitioned into 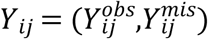 and 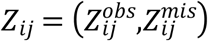, respectively.

Joint model given the random effects is given as

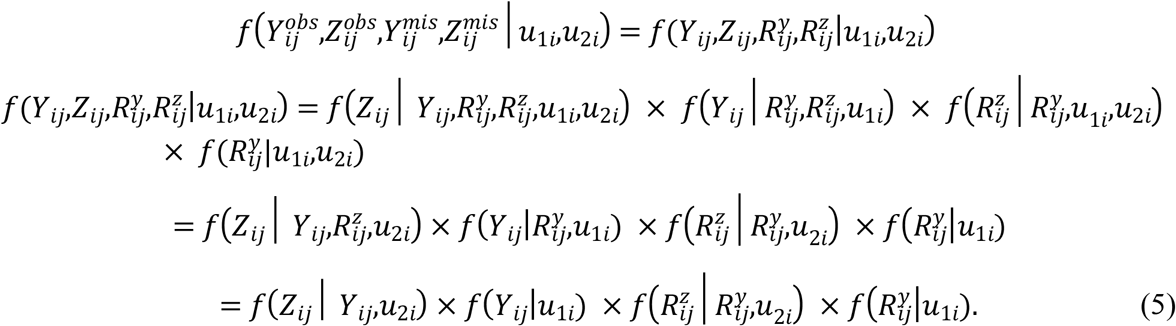

Joint probability distribution function is formulated further as

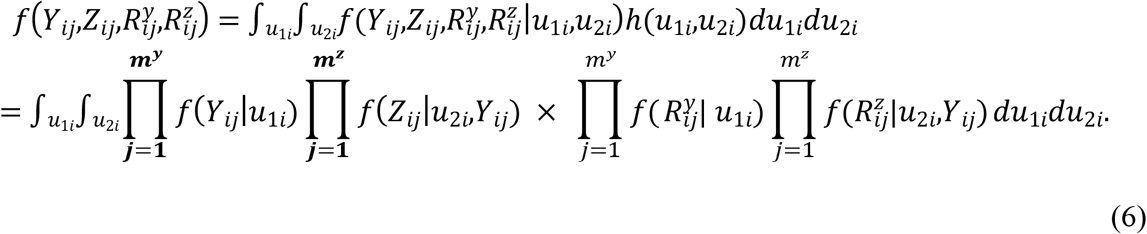

### 3.3 Prior distributions and hierarchical model

The unknown parameters of the proposed model are estimated by employing the Bayesian framework using Markov chain Monte Carlo (MCMC) techniques [18]. MCMC has an advantage over conventional methods, with MCMC conditional distribution of each parameter given others is easily specifying. Priors are chosen to carry out Bayesian inference for unknown parameters, let *Ø* = {*α*_1_,*α*_2_,*σ*^2^,*D*_1_,*D*_2_,*Φ*_1_,*Φ*_2_,*π*^*y*^,*π*^*z*^,*ϒ*_*l*_,*κ*_*k*_} be the unknown parameters’ vector where *k* = 1,2,3,…., *m*^*y*^ and *l* = 1,2,3,….,*m*^*z*^. We specify proper prior for each set of unknown parameters: Gaussian distribution for the fixed effects, and Inverse Gamma distribution for random effects. Different levels of variances for each distribution are tried out to make robust choice of fixed effects estimates. Independent prior distributions for unknown parameters are chosen by assigning hyperparameters that lead to low-informative prior distributions.

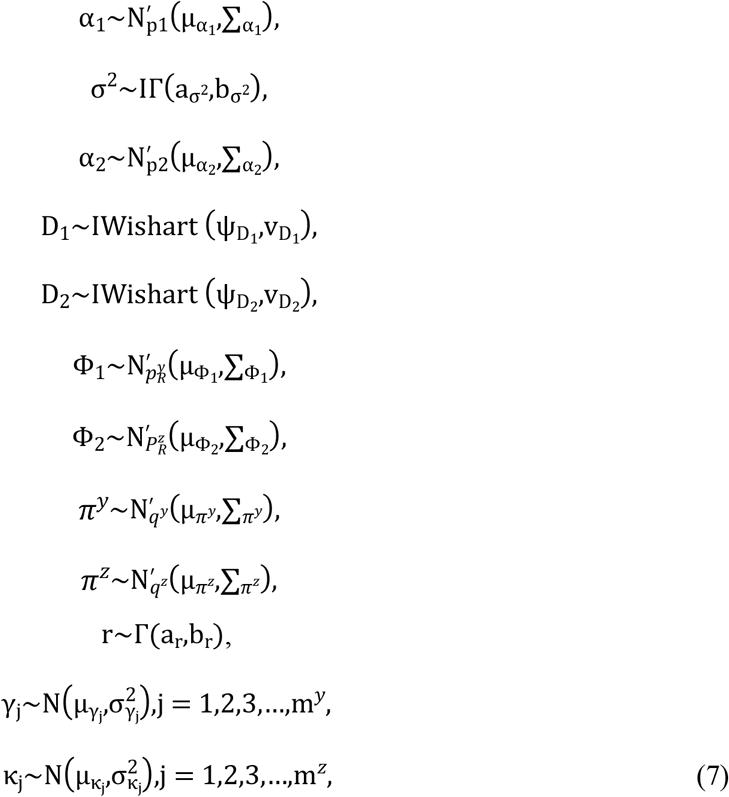

where IΓ(a,b) and Γ(a,b) denote inverse gamma distribution and gamma distribution with shape parameter a and scale parameter b. IW*i*shart (ψ, v) represents the inverse Wishart distribution with scale and matrix parameters v and ψ, respectively. N(μ, ∑) denotes a normal distribution with mean vector μ and covariance matrix ∑.

The joint posterior density is formulated as

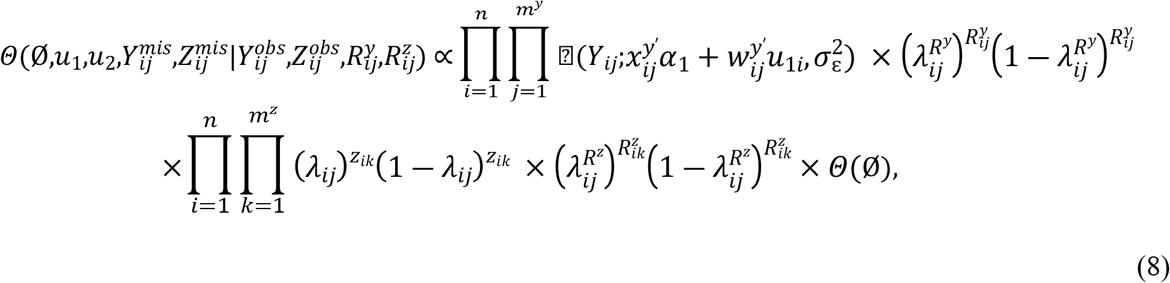

where, *Θ*(*Ø*) is the joint prior distribution of unknown parameters.

Samples are drawn iteratively from conditional posterior distributions derived from (8) using Gibbs sampler, the full conditional distributions for parameters are given by

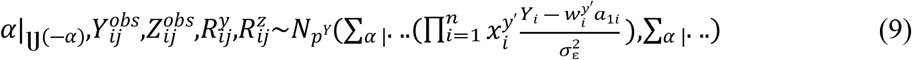

Where 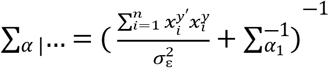

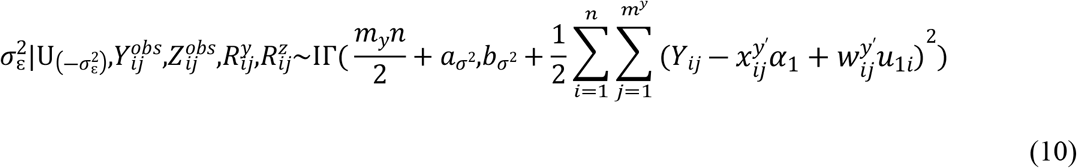

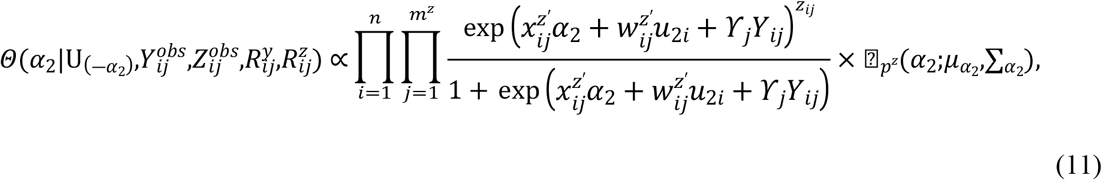

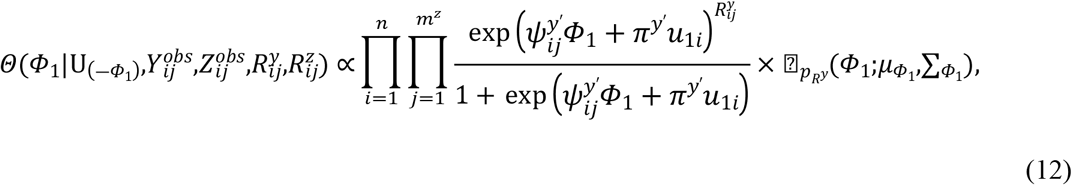

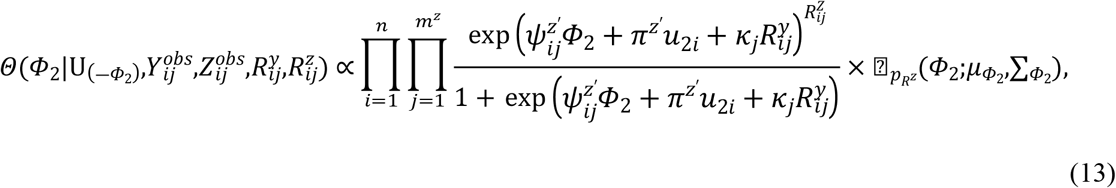

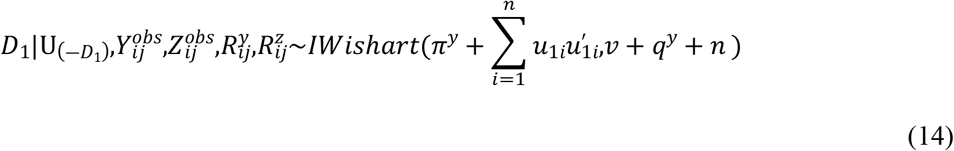

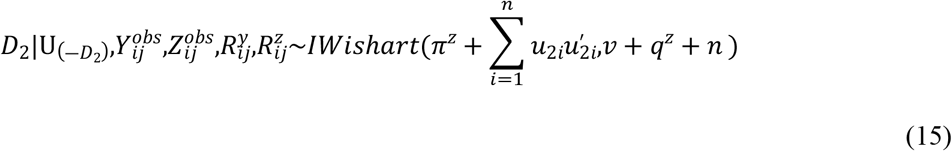

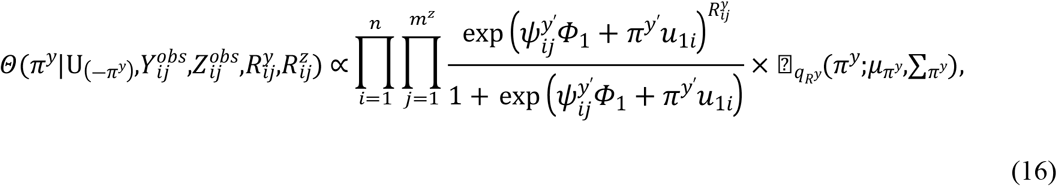

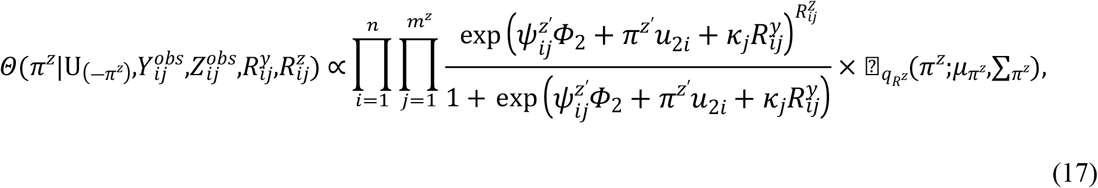

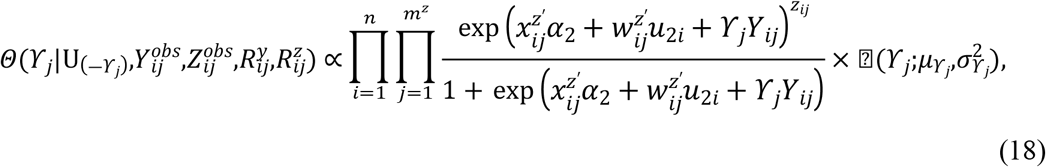

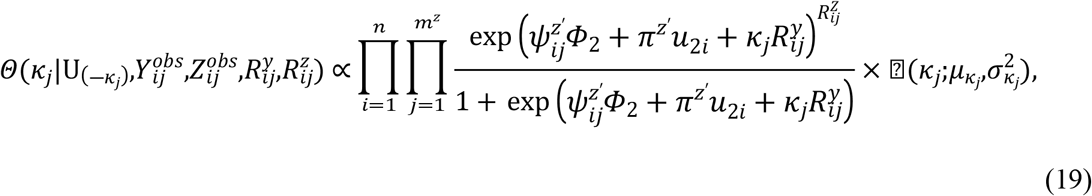

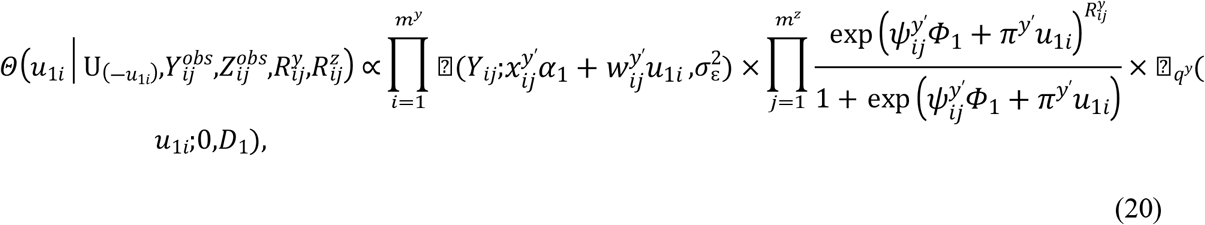

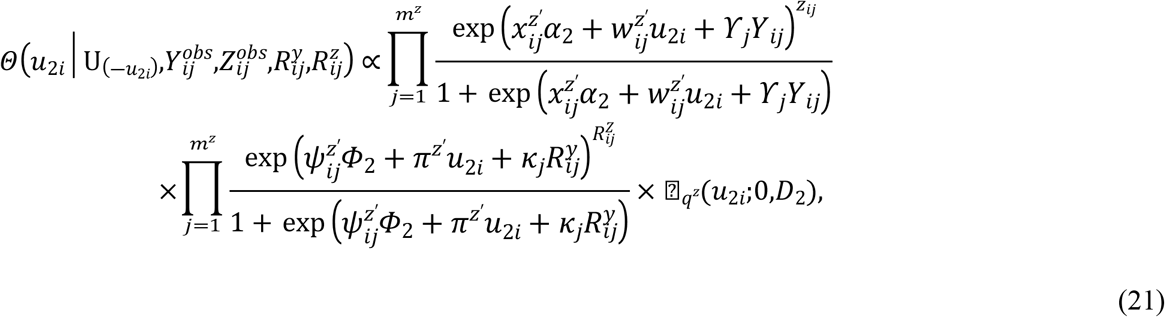

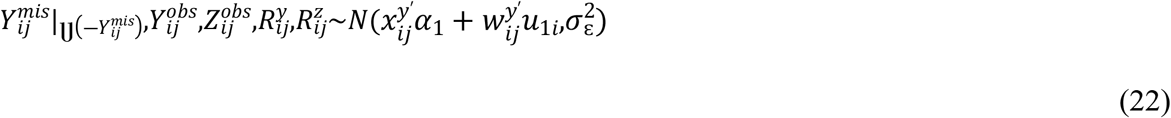

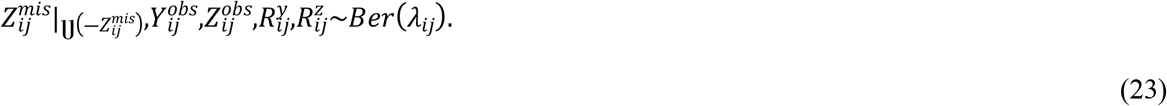

## 4. Simulation studies

Some simulation studies are conducted to assess the performance of the proposed model. The first simulation is designed with assuming no association between two responses, and between measurements and missingness processes. The second simulation assumes different associations between biomarkers, also between biomarkers and missingness processes. Each simulation has 100 random samples and each sample includes 500, 1,000, and 3,000 subjects.

For each individual, the two longitudinal measurements are generated from the following joint model,

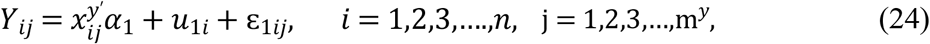

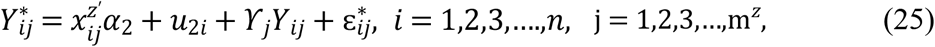

where 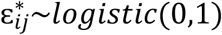, *Z*_*ij*_ = 1 for 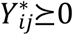 and *Z*_*ij*_ = 0 for 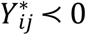

For missingness processes, following models are assumed to generate data,

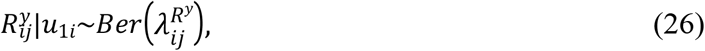

where, 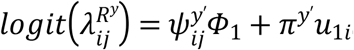

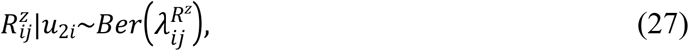

where, 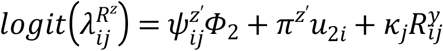

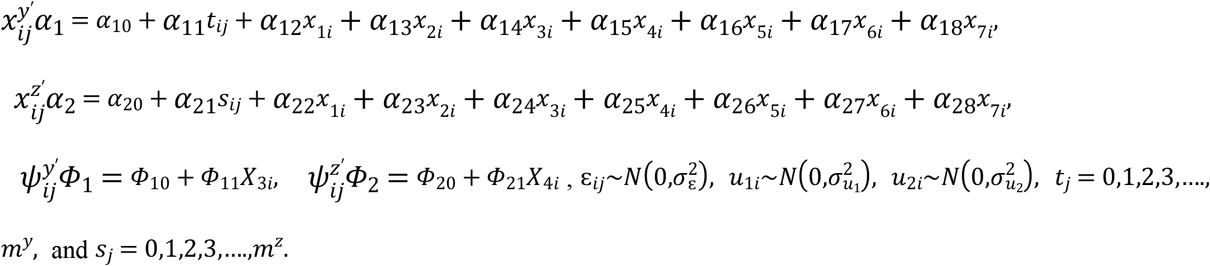

To make simulation feasible, each dataset included small to large individuals with longitudinal measurements. Follow-up visits are scheduled at time = {0, 1, 2, 3, 4, 5}. The true values considered for *α*_1_ = *α*_2_ = (1, ― 1,0,2,1,1,1, ― 1, ― 1)′ and *Φ*_1_ = *Φ*_2_ = (2,1)′. *x*_11_,*x*_12_,*x*_13_,*x*_14_ are generated from *N*(0,1), and *x*_15_,*x*_16_,*x*_17_ are generated from *binom*(*n*,1,0.1). *u*_*i*_ = (*u*_1*i*_,*u*_2*i*_) is generated from *MVN* = (0,*D*), where

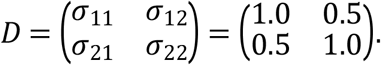

For the SPM incorporating non-monotone, non-ignorable missing data mechanism, model (26) and model (27) are formulated following Fitzmaurice et al. [19]. We first considered independence (*ϒ*_*j*_,*κ*_*j*_) = (0,0)′. Sample sizes are assessed by considering *n* = 500, *n* = 1,000, and *n* = 3,000 individuals with different association parameters as (*ϒ*_*j*_,*κ*_*j*_) = (1.000, ― 1.000)′, (*ϒ*_*j*_,*κ*_*j*_) = (0.500, ― 0.500)′, (*ϒ*_*j*_,*κ*_*j*_) = (― 0.500, ― 0.500)′.

Sensitivity analysis of posterior distribution is assessed by selecting different prior distributions. In the case of informative priors, the structure of prior distributions take main focus during the sensitivity analysis. Non-informative priors are assessed based on changes in posterior inference. All our parameters are checked for different parameters’ values to assess robustness of posterior means. In addition, sensitivity analysis can also be done for missingness mechanisms, where the assessment for ignorable and non-ignorable missingness can be evaluated. We do not apply sensitivity analysis for different missingness mechanisms, as it is beyond the scope of our study objective.

A computational procedure to estimate parameters in proposed joint model is conducted using Gibbs sampling by WinBUGS software. Two parallel MCMC sampling chains run with different starting values, convergence of chains is examined by trace plots and with diagnostic statistics suggested by Gelman et al. [18]. Posterior estimates are based on 10,000 iterations after discarding 5,000 of the burn-in period.

Data are generated under two scenarios: biomarker variability is not associated with each other (*ϒ* = 0), and both biomarkers are associated with each other (*ϒ* = ± 0.5, *or* ± 1.0). With that, it is also evaluated whether either missingness process is associated with the measurements process (*κ* = ± 0.5, 0*r* ± 1.0) or not. Each dataset is then analyzed using our proposed joint model.

Based on the results presented in Table 1, it is to be noted that the performance of the proposed joint model is good. Consistency of parameter estimates is evaluated by increasing sample size which causes reduced bias and standard error for respective parameters. The same interpretation of parameter estimates holds for zero to non-zero association parameters.

**Table 1:** Results of the simulation study for binary and continuous longitudinal biomarkers data incorporating NMAR based on joint modelling for the simulation study. Estimate (Est.), standard error (SE), Bias for *N* = 100 simulated data with sample size 500, 1,000, and 3,000 considering association (*ϒ*_*j*_,*κ*_*j*_) = (0.5, ― 0.5)′.

| Parameter | Real | *n=500* |  | *n=1,000* |  | *n=3,000* |  |
| --- | --- | --- | --- | --- | --- | --- | --- |
|  |  | Est(SE) | Bias | Est(SE) | Bias | Est(SE) | Bias |
| $\alpha_{10}$ | 1.000 | 1.130(0.260) | 0.130 | 0.895(0.161) | -0.105 | 1.088(0.101) | 0.088 |
| $\alpha_{11}$ | -1.000 | -1.142(0.196) | -0.142 | -0.985(0.119) | 0.015 | -1.006(0.075) | -0.006 |
| $\alpha_{12}$ | 0.000 | 0.275(0.118) | 0.275 | 0.112(0.073) | 0.112 | -0.062(0.043) | -0.062 |
| $\alpha_{13}$ | 2.000 | 2.587(0.397) | 0.587 | 1.896(0.242) | -0.104 | 1.969(0.152) | -0.031 |
| $\alpha_{14}$ | 1.000 | 1.066(0.209) | 0.066 | 0.943(0.135) | -0.057 | 0.971(0.082) | -0.029 |
| $\alpha_{15}$ | 1.000 | 1.093(0.215) | 0.093 | 1.028(0.139) | 0.028 | 1.016(0.086) | 0.016 |
| $\alpha_{16}$ | 1.000 | 1.133(0.366) | 0.133 | 1.586(0.257) | 0.586 | 0.962(0.151) | -0.038 |
| $\alpha_{17}$ | -1.000 | -0.622(0.402) | 0.378 | -1.121(0.288) | -0.121 | -0.991(0.162) | 0.009 |
| $\alpha_{18}$ | -1.000 | -1.169(0.376) | -0.169 | -0.863(0.264) | 0.137 | -0.926(0.168) | 0.074 |
| $\alpha_{20}$ | 1.000 | 1.056(0.062) | 0.056 | 1.050(0.043) | 0.050 | 1.044(0.025) | 0.044 |
| $\alpha_{21}$ | -1.000 | -1.007(0.005) | -0.007 | -0.998(0.004) | 0.002 | -1.000(0.002) | 0.000 |
| $\alpha_{22}$ | 0.000 | -0.045(0.021) | -0.045 | -0.033(0.030) | -0.033 | -0.005(0.002) | -0.005 |
| $\alpha_{23}$ | 2.000 | 1.976(0.039) | -0.024 | 1.993(0.028) | -0.007 | 1.999(0.015) | -0.001 |
| $\alpha_{24}$ | 1.000 | 0.978(0.037) | -0.022 | 0.980(0.029) | -0.020 | 1.007(0.015) | 0.007 |
| $\alpha_{25}$ | 1.000 | 1.036(0.039) | 0.036 | 1.029(0.027) | 0.029 | 1.006(0.016) | 0.006 |
| $\alpha_{26}$ | 1.000 | 1.076(0.122) | 0.076 | 1.069(0.092) | 0.069 | 1.022(0.053) | 0.022 |
| $\alpha_{27}$ | -1.000 | -1.102(0.132) | -0.102 | -1.040(0.095) | -0.040 | -1.007(0.050) | -0.007 |
| $\alpha_{28}$ | -1.000 | -0.765(0.120) | 0.235 | -1.159(0.087) | -0.159 | -1.020(0.051) | -0.020 |
| $\Phi_{10}$ | 2.000 | 1.845(0.136) | -0.155 | 1.992(0.101) | -0.008 | 1.997(0.060) | -0.003 |
| $\Phi_{11}$ | 1.000 | 1.039(0.077) | 0.039 | 1.030(0.058) | 0.030 | 1.008(0.033) | 0.008 |
| $\Phi_{20}$ | 2.000 | 1.967(0.080) | -0.031 | 1.989(0.058) | -0.011 | 1.991(0.034) | -0.009 |
| $\Phi_{21}$ | 1.000 | 0.973(0.078) | -0.027 | 0.990(0.056) | -0.010 | 0.999(0.032) | -0.001 |
| $\sigma_{\varepsilon}^2$ | 1.000 | 1.029(0.029) | 0.029 | 1.023(0.020) | 0.023 | 1.006(0.012) | 0.006 |
| $\sigma_{u_1}^2$ | 1.000 | 0.884(0.074) | -0.116 | 0.902(0.056) | -0.098 | 0.989(0.031) | -0.011 |
| $\sigma_{u_2}^2$ | 1.000 | 1.562(0.556) | 0.562 | 1.193(0.284) | 0.193 | 1.140(0.191) | 0.140 |
| $Y_1$ | 0.500 | 0.341(0.189) | -0.159 | 0.471(0.118) | -0.029 | 0.495(0.075) | -0.005 |
| $Y_2$ | 0.500 | 0.276(0.187) | -0.224 | 0.428(0.119) | -0.072 | 0.502(0.076) | 0.002 |
| $Y_3$ | 0.500 | 0.614(0.239) | 0.114 | 0.559(0.178) | 0.059 | 0.521(0.084) | 0.021 |
| $Y_4$ | 0.500 | 0.534(0.249) | 0.034 | 0.526(0.188) | 0.026 | 0.512(0.101) | 0.012 |
| $Y_5$ | 0.500 | 0.538(0.425) | 0.038 | 0.532(0.224) | 0.032 | 0.509(0.116) | 0.009 |
| $Y_6$ | 0.500 | 0.603(0.353) | 0.103 | 0.554(0.283) | 0.054 | 0.513(0.353) | 0.013 |
| $\kappa_1$ | -0.500 | -0.562(0.177) | -0.062 | -0.556(0.134) | -0.056 | -0.517(0.077) | -0.017 |
| $\kappa_2$ | -0.500 | -0.351(0.182) | 0.149 | -0.442(0.131) | 0.058 | -0.490(0.078) | 0.010 |
| $\kappa_3$ | -0.500 | -0.428(0.189) | 0.072 | -0.449(0.132) | 0.051 | -0.496(0.076) | 0.004 |
| $\kappa_4$ | -0.500 | -0.441(0.180) | 0.059 | -0.450(0.133) | 0.050 | -0.498(0.077) | 0.002 |
| $\kappa_5$ | -0.500 | -0.514(0.181) | -0.014 | -0.510(0.132) | -0.010 | -0.506(0.078) | -0.006 |
| $\kappa_6$ | -0.500 | -0.521(0.185) | -0.021 | -0.516(0.132) | -0.016 | -0.507(0.078) | -0.007 |
| $\pi^y$ | 1.000 | 1.063(0.043) | 0.063 | 0.998(0.035) | -0.002 | 0.998(0.029) | -0.002 |
| $\pi^z$ | 1.000 | 1.183(0.711) | 0.183 | 1.058(0.127) | 0.058 | 1.037(0.118) | 0.037 |

## 5. Prostate cancer data analysis

The primary objective of prostate cancer (PC) data analysis is to detect any potential association between PSA and ALP biomarkers, and to simultaneously analyze both. PC data were collected from *n* =1504 patients who had at least 2 measurements of PSA, and ALP. Patients’ *Age, Platelets, BMI, Bilirubin, Gleason Score, Grade* are recorded along with the prescribed *Drug*. Following joint model is considered,

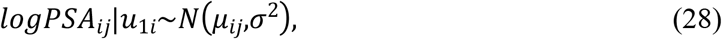

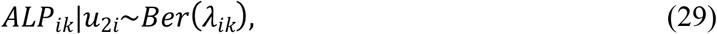

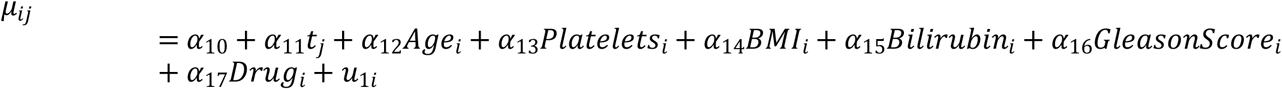

and

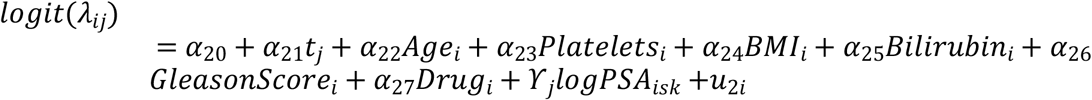

In addition, for non-ignorable missingness mechanisms following models are proposed,

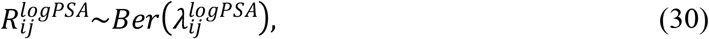

where 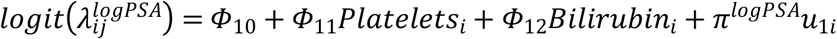

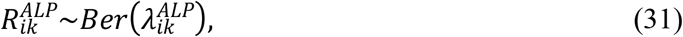

where, 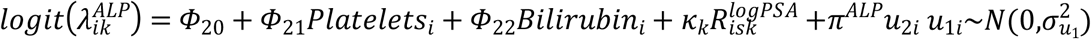, and 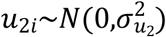

Two parallel MCMC chains runs with different initial values for 10,000 iterations and discarded first 5,000 iterations as pre-convergence burn-in. Convergence of MCMC chains is checked using Gelman-Rubin diagnostic test. In addition, trace plots can plot for unknown parameters to assess convergence. To get posterior inference, prior distributions for unknown parameters are selected as, u_i_ = (u_1i_,u_2i_)∼MVN((0,0),D_u_), *i* = 1,…,*n*, where D_u_∼IWishart 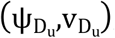, such that the hyper-parameters of 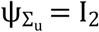 and 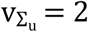 which lead to low-informative priors. It is also to be assumed that 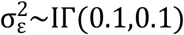, the regression coefficients (*α*_*i*1_,*α*_*i*2_,*ψ*_*i*1_,*ψ*_*i*2_)′ are fixed effects unknown parameters and the prior distributions for them are N(0,1000), γ_j_, *j* = 1,…,*m*^*y*^ is the associated parameter of the continuous longitudinal log(PSA) on the binary longitudinal ALP at time j and the prior distributions for it is N(0,1000). Prior distribution for associated parameter κ_j_, *j* = 1,…,*m*^*z*^ is N(0,1000), *π*^*log****P****SA*^,*π*^*AL****P***^∼*N*(0,1000).

For model selection, we apply the deviance information criterion (DIC), which identifies the most suitable model by balancing between likelihood and parameters’ numbers. Posterior means and the number of parameters are used in this criteria to find the best-fitted model [20].

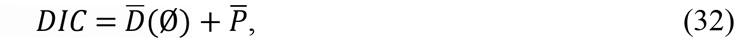

Where, 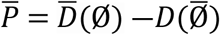 and 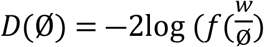.

*D*(*Ø*) and 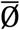 are posterior estimates of *D*(*Ø*) and *Ø. w* = *w*_*i*_ = (*w*_1_,*w*_2_,*w*_3_,….,*w*_*n*_) is full data with marginal density, 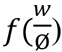 and *Ø* are vectors of model parameters. The lowest DIC leads us to the best model fit, that is our proposed joint model.

The parameter estimates and their 95% posterior intervals are presented in Table 2 and summarized for joint model: PSA level decreases with respect to time after treatment, and it is a good sign to evaluate efficiency of prescribed treatment when this decrease is significant. PSA alone is not an adequate biomarker, ALP measurements are also taken into account. ALP increases with respect to time for individuals, but this increase has not had any significant effect. The results indicate that with increasing age, both PSA and ALP increase. This positive increase is significant for PSA but non-significant for ALP. Platelets are another predictor of increasing PSA significantly, but due to one unit increase in platelets, ALP level decreases non-significantly with 0.026 on average. With increasing BMI, patients’ PSA and ALP both tend to significantly decrease. One unit increase in bilirubin measurement makes a significant increase of 0.201, and 0.617 in PSA and ALP measurements, respectively. The results show that PSA and ALP measurements are lower among those patients with a Gleason score greater than or equal to (4+3) as compared to those whose score is lower than and equal to (3+4), it is due to treatment effect. This study’s results revealed that patients who received ALT, prostatectomy, and their combinations have lower PSA and ALP measurements as compared to those who received EBRT; and this is a good sign to prove drug efficacy. There exists a positive correlation between PSA and ALP measurements that can be revealed with associated parameters *ϒ*; while a negative association exists between missingness and measurement processes. Results revealed that due to the existence of a positive association both PSA and ALP measurements should be simultaneously taken on PC patients to get insights about PC progression. In addition, missingness must be incorporated into data to avoid loss of information.

**Table 2:**
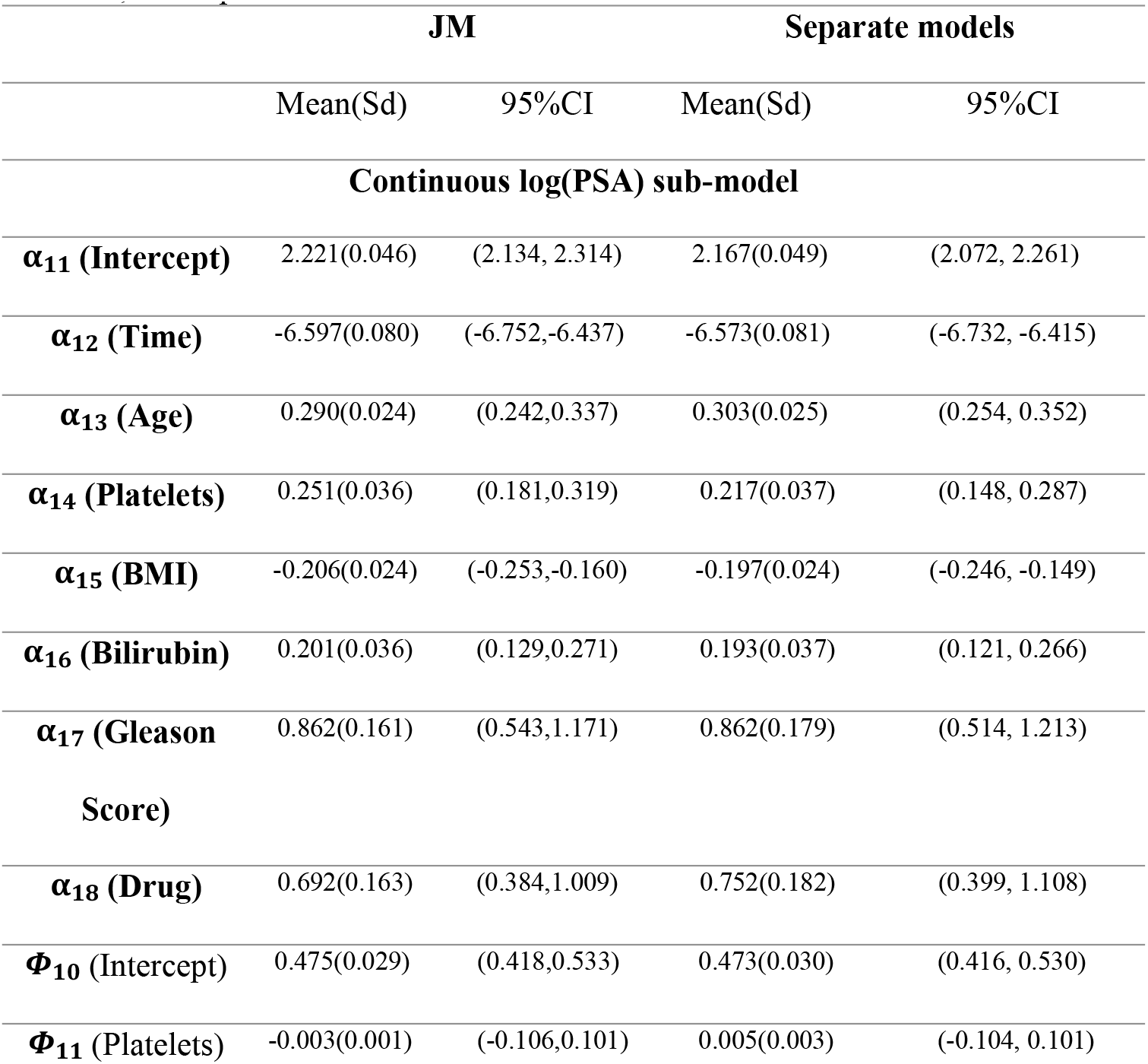
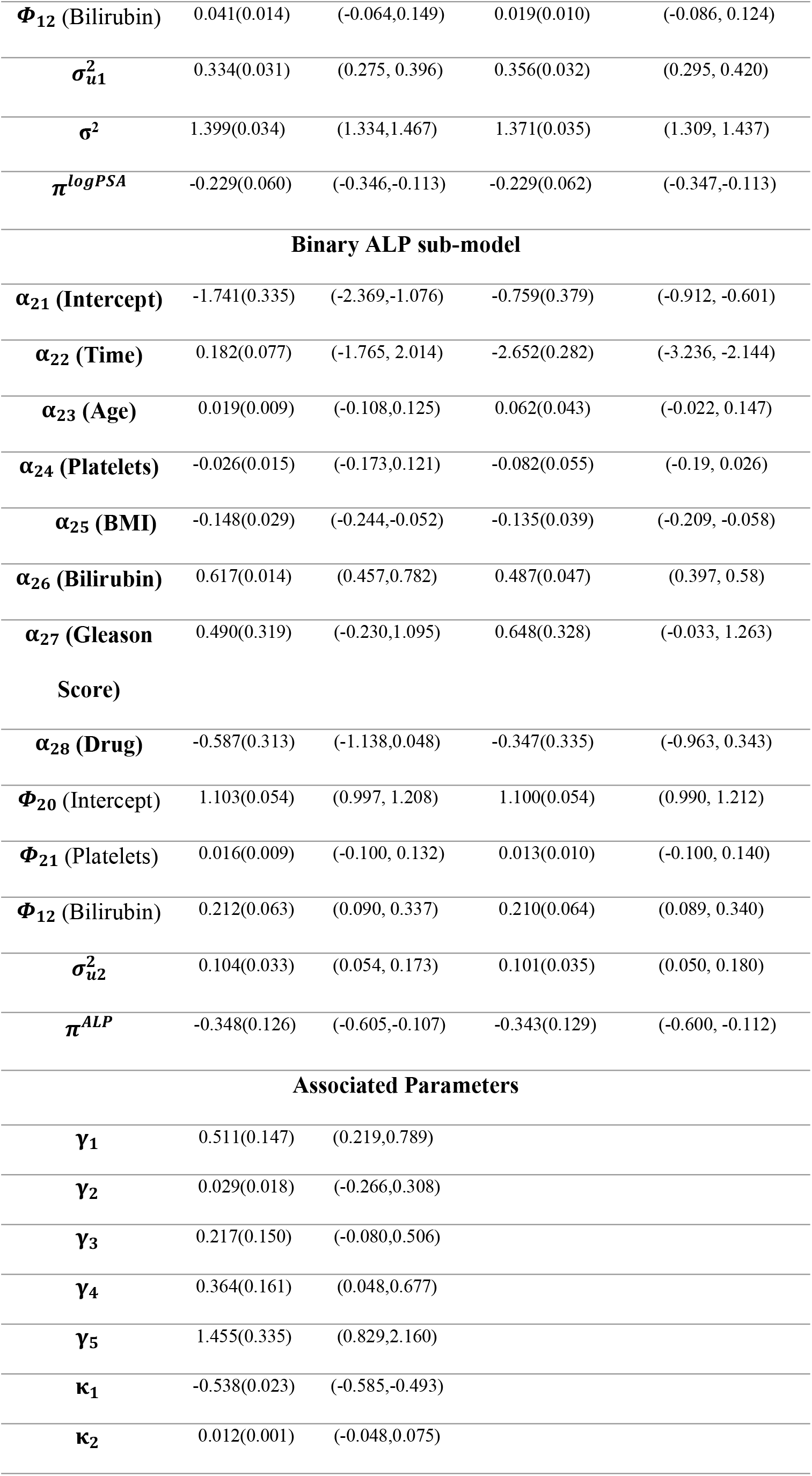
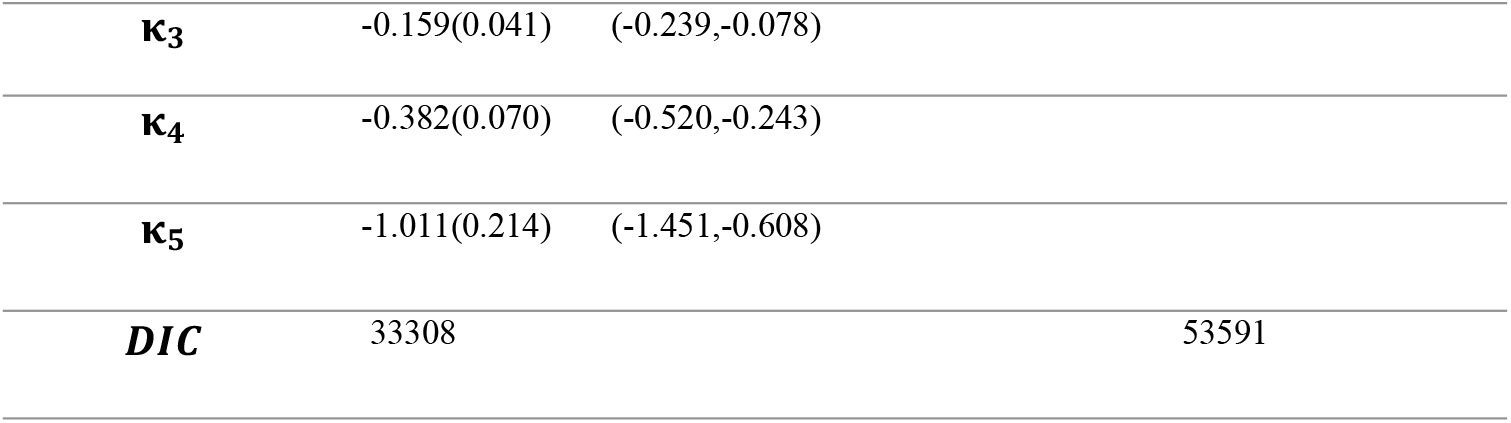
Parameter estimates (Mean), standard deviation (Sd), and 95% CI (credible interval) of PC data, by applying our proposed joint SPM, and separate models.

## 6. Conclusion

The primary goal of this manuscript was to jointly model different types of longitudinal outcomes incorporating non-ignorable missingness. One can directly apply mixed modelling techniques with considering missingness as an ignorable phenomenon. However, follow-up studies could get over the hurdles of loss of information in case of missingness by proposing SPM for mixed-longitudinal outcomes with missingness. For each missing data process, a logit model is proposed via a latent variable to depict the tendency of change for individuals. SPM proposed to join longitudinal and missingness models using random effects. This paper presents a joint modelling approach to analyze longitudinal biomarkers measurements in the presence of non-ignorable missing data due to both intermittent and monotone missingness. In our PC dataset, the non-monotone missingness pattern exists due to patients who missed visits, and monotone missingness data are from those patients who left the follow-up and never came back to complete the treatment process. We use logistic models to describe the missingness patterns. Our proposed model is adequate to account for the association between measurements and missingness processes, and it can be extended to models with more than two mixed longitudinal outcomes incorporating more than two missing outcomes processes.

Correlated random effects provide inference for non-ignorable missing data. However, the non-ignorable missingness assumption is untestable for the data at hand. In our collected PC dataset, non-ignorable assumption is verified as missingness occurred due to lack of treatment efficacy. Therefore, we incorporated missingness process in data analysis. However, researchers must put extra care to check assumptions about missingness mechanism, if the cause of missing data is not internally related to responses; local sensitivity analysis should be performed.

This study emphasis that PC patients must be monitored for PSA and ALP simultaneously. It is important to take into consideration both PSA and ALP levels as both can influence the health of PC patients. In addition, missing observations must be incorporated in data analysis to get full information about patients’ health and wellbeing. An elevated level of PSA shows the non-effectiveness of particular treatment, and an elevated level of ALP depicts the spread of PC tumor cells.

## Data Availability

Data cannot be shared publicly because it is secondary data taken from a hospital. Data is available from the corresponding author upon reasonable request for researchers who meet the criteria for access to confidential data.

## Acknowledgements

None.

